# Towards definitions of critical illness and critical care using concept analysis

**DOI:** 10.1101/2022.01.09.22268917

**Authors:** Raphael Kazidule Kayambankadzanja, Carl Otto Schell, Martin Gerdin Wärnberg, Thomas Tamras, Hedi Mollazadegan, Mats Holmberg, Helle Mølsted Alvesson, Tim Baker

## Abstract

**Objective:** As “critical illness” and “critical care” lack consensus definitions, this study aims to explore how the concepts’ are used, describe their defining attributes and propose potential definitions.

**Design:** We used the Walker and Avant stepwise approach to concept analysis. The uses and definitions of the concepts were identified through a scoping review of the literature and an online survey of 114 global clinical experts. Through content analysis of the data we extracted codes, categories and themes to determine the concepts’ defining attributes and we proposed potential definitions. To assist understanding, we present model, related and contrary cases concerning the concepts, we identified antecedents and consequences to the concepts, and defined empirical referents.

**Results:** The defining attributes of critical illness were a high risk of imminent death; vital organ dysfunction; requirement for care to avoid death; and potential reversibility. The defining attributes of critical care were the identification, monitoring and treatment of critical illness; vital organ support; initial and sustained care; any care of critical illness; and specialized human and physical resources. Our proposed definition of critical illness is, “*a state of ill health with vital organ dysfunction, a high risk of imminent death if care is not provided and the potential for reversibility*”. Our proposed definition of critical care is, “*the identification, monitoring and treatment of patients with critical illness through the initial and sustained support of vital organ functions*.”

**Conclusion:** The concepts critical illness and critical care lack consensus definitions and have varied uses. Through concept analysis of uses and definitions in the literature and among experts we have identified the defining attributes of the concepts and propose definitions that could aid clinical practice, research, and policy making.

**Strengths and Limitations of the Study:**

- This concept analysis is the first study to systematically describe the uses and definitions of the concepts *critical illness* and *critical care*
- The study uses a scoping review of the literature and input from over one hundred clinical experts from diverse settings globally to identify the defining attributes and provide proposed definitions of the concepts
- Some uses and definitions of the concepts in languages other than English, in unpublished grey literature and from clinical experts not included in the study may have been missed
- As current usage of the concepts is diverse, the proposed definitions may not be universally accepted and are aimed to stimulate further discussion

## Introduction

The concepts *critical illness* and *critical care* are commonly used in healthcare. In PubMed, both are Medical Subject Headings (MeSH) terms, and searches for “critical illness” or “critical care” return 40,000 and 220,000 articles respectively. While it may seem evident that the concepts concern patients with very serious illness and their care, there is a lack of consensus around their precise definitions.

This causes problems for clinical practice, research, and policy making. For the clinician, discordant interpretations of when a patient is critically ill can lead to differing clinical assessments and treatments despite similar states: when should a patient be regarded as critically ill so that an alarm should be triggered and when is admission to an intensive care unit warranted? For the researcher, it can be difficult to design a study or interpret findings: studying the effect of a treatment for critical illness requires clear eligibility criteria and translating the findings to another patient group requires that the groups have similar clinical conditions. For the policy maker, prioritising programmes and investments designed to improve care for very sick patients relies on comparisons between similar groups and clearly defined interventions.

Even quantifying the total global burden of critical illness has been challenging due to the lack of an agreed definition. Proxies have been used instead, for example summing up syndromes considered to comprise critical illness such as sepsis and acute lung injury– resulting in estimates of up to 45 million critical illness cases each year.(1) Low- and middle-income countries are suspected to have the highest burden (2), but the lack of a definition has hampered comparisons across settings.

Studying the care for critically ill patients has also been problematic. Studies have focused on care provided in hospital locations such as in intensive care or emergency units, which exclude care provided in hospitals lacking such units, and to critically ill patients in general hospital wards. (3–5) In the COVID-19 pandemic, there have been great efforts to describe, scale-up and improve care for critically ill patients throughout the world (3,5) and a lack of agreement around critical care has hampered these efforts.

These examples illustrate how important concepts are as the building blocks of theories and communication. Ideally, concepts are clearly defined and their use well described for unambiguous communication and an understanding about exactly what is being described or explained. (6) *Concept analysis* is a method for investigating how concepts are used and understood. Concept analyses have been conducted in diverse fields such as in teamwork (7), postoperative recovery(8) and bioterrorism preparedness(9), all with the aim of providing basic conceptual understanding and facilitating communication. In this paper, we have used concept analysis, following the stepwise approach described by Walker and Avant(6). The first two steps in the approach are to choose the concept and determine the aim of the analysis. Our chosen concepts are *critical illness* and *critical care* and our aims are to explore the uses and definitions of the concepts in published sources and by global clinical experts, leading to a description of the defining attributes of the concepts and to proposed definitions.

## Methods

The Walker & Avant approach to concept analysis uses the following steps: identifying the uses of the concept; determining the concept’s defining attributes; presenting a model case, identifying related and contrary cases; identifying antecedents and consequences; and defining empirical referents.(6)

### Identifying the uses of the concept

We identified the uses of the concepts of critical illness and critical care through a scoping review of the literature and a web-based survey of global experts.

#### Scoping Review

We used the Arksey and O’Malley framework for scoping reviews(10). Relevant studies in English were identified in the PubMed and Web of Science databases. To include publications that were not found through the database searches, hand-searching of publication lists of intensive care medicine, and emergency medicine societies was performed. Duplicates were removed using the online software program Rayyan(11). The publications were examined through title, then abstract review and lastly by full-text review.

##### Critical Illness

The search terms used were terminolog*, etymolog*, nomenclatur*, definition*, plus emergency, critical*, acute*, sever*, ill, illness. A total of 9323 articles were identified using these critical illness terms in the databases and an additional two articles were identified through hand-searching. Of these, 1126 articles were identified as duplicates and the remaining 8199 articles were screened by title and abstract review by two of the authors (TT and HM). 8168 articles were excluded as they did not concern critical illness, were not written in English or were not available in full text online, leaving 31 articles for inclusion for full-text review. In the full-text review, 22 articles were excluded as they did not define critical illness, and so nine articles were included in the analysis (Supplementary Table 1).

##### Critical Care

The search terms used were terminolog*, etymolog*, nomenclatur*, definition*, plus critical care, intensive care, emergency care, acute care. A total of 7286 articles were identified using these critical care terms in the databases and an additional six articles were identified through hand-searching. Of these, 1964 were identified as duplicates and the remaining 5322 articles were screened by title and abstract review by two of the authors (TT and HM). 5269 articles were excluded as they were not concerning critical illness, not written in English or not available in full text online, leaving 59 articles for inclusion for full-text review. In the full-text review, 46 articles were excluded as they did not define critical care and so 13 articles were included in the analysis (Supplementary Table 2).

#### Expert survey

The survey used open-ended questions to gather information about the experts’ definitions of critical illness and critical care, and how they see the relationship of the concepts to connected concepts in order to provide context. The survey included the questions: i. *How would you define critical illness*?, ii. *How would you define critical care?*, iii. *Do critical care and intensive care differ*? *If yes, in what way?* iv. *Do critical care and emergency care differ and if yes, in what way?* v. *Do critical care and acute care differ and if yes, in what way*?

The inclusion criterion for an expert to be invited to participate in the survey was experience in any medical specialty that includes care of patients with acute, severe illness. Experts were identified from a stakeholder mapping of global critical care done by one of the authors (TB, unpublished), and those known to the researchers to be global experts in the field of critical care. Purposive sampling was used to invite experts with the aim of including 100 experts with a balance between specialties, geographical locations, health worker cadres and gender. In total 146 experts were invited to take part, and 113 completed the survey (77% response rate) (Table 1).

**Table 1:** Characteristics of the experts who participated in the survey.

| Variable | Frequency (%) |
| --- | --- |
| All | 114 |
| Gender |  |
| Male | 80 (70.2) |
| Female | 34 (29.8) |
| Continent |  |
| Africa | 42 (36.8) |
| Europe | 29 (25.4) |
| North America | 26 (22.8) |
| Asia | 12 (10.5) |
| South America | 3 (2.6) |
| Australia | 2 (1.8) |
| Cadres* |  |
| Physician | 93 (53.1) |
| Researcher | 62 (35.4) |
| Nurse | 12 (6.9) |
| Policy Maker | 5 (2.9) |
| Other | 3 (1.7) |
| Specialty* |  |
| Anaesthesia/Intensive Care | 75 (59.1) |
| Emergency Care | 20 (15.8) |
| Medicine | 12 (9.5) |
| Paediatrics | 7 (5.5) |
| Surgery | 6 (4.7) |
| Obstetrics and Gynaecology | 2 (1.6) |
| Other | 5 (3.9) |

\* As the experts were asked to select all that apply, the sum may exceed 100%

### Analysis and determining the defining attributes

The definitions of critical illness and critical care from the scoping reviews and the expert survey were charted and analysed using a content analysis based on methods developed by Erlingsson & Brysiewicz.(12) First, the data from any parts of the literature and from the expert survey that concerned the uses or definitions of the concepts were extracted. The data were coded and the codes analysed iteratively by the study team. Redundant codes were removed and similar codes were arranged into categories. The data were revisited when new categories arose or when diverse opinions with contrasting attributes were identified. Through the process, themes emerged that captured the defining attributes of the concepts. Using the defining attributes, definitions of the concepts were constructed by the research team to be coherent and useful.

### Presenting a model case, related and contrary cases, identifying antecedents and consequences, and defining empirical referents

The model cases, related, and contrary cases were developed by the researchers to provide examples to illustrate the defining attributes of the concepts that emerged from the concept analysis. Model cases were developed to be clinically realistic and to include all the defining attributes. Related cases were developed that include some, but not all, of the defining attributes, and contrary cases that are clearly “not the concept”, containing none of the defining attributes. For simplicity in this study, we limited our cases to examples of patients with respiratory disease. Antecedents and consequences were identified as events that occur prior to the occurrence of each concept and as the outcomes of each concept respectively. Empirical referents were identified as phenomena that demonstrate the occurrence of each concept “in real life”.

#### Ethical considerations

Informed consent was provided by all of the experts. The Research Ethics Committee of the London School of Hygiene and Tropical Medicine approved the study (Reference number 22661).

## Results

The results relate to steps 4-8 in the Walker and Avant approach, as steps 1-3 have been described in the introduction and methods.

### Critical Illness

#### Defining attributes

A total of 48 codes were identified from the uses and definitions of critical illness from the scoping review and expert survey. The codes were analysed into 14 categories and 4 themes. (Table 2). The themes represent the defining attributes of critical illness: *high risk of imminent death*; *vital organ dysfunction*; *requirement for care to avoid death;* and *potential reversibility*. (Figure 1)

**Table 2.**
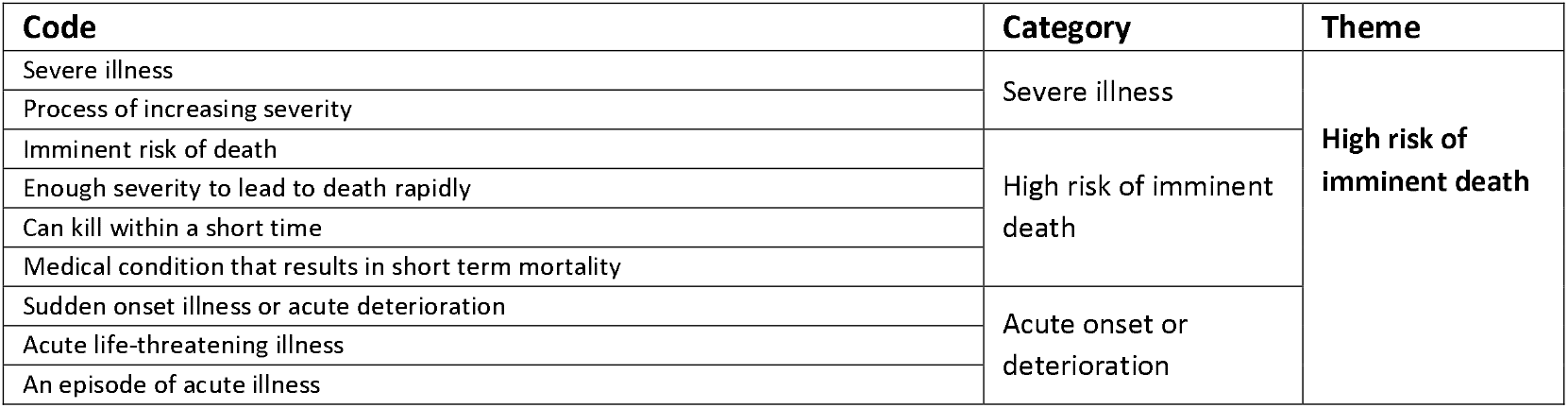
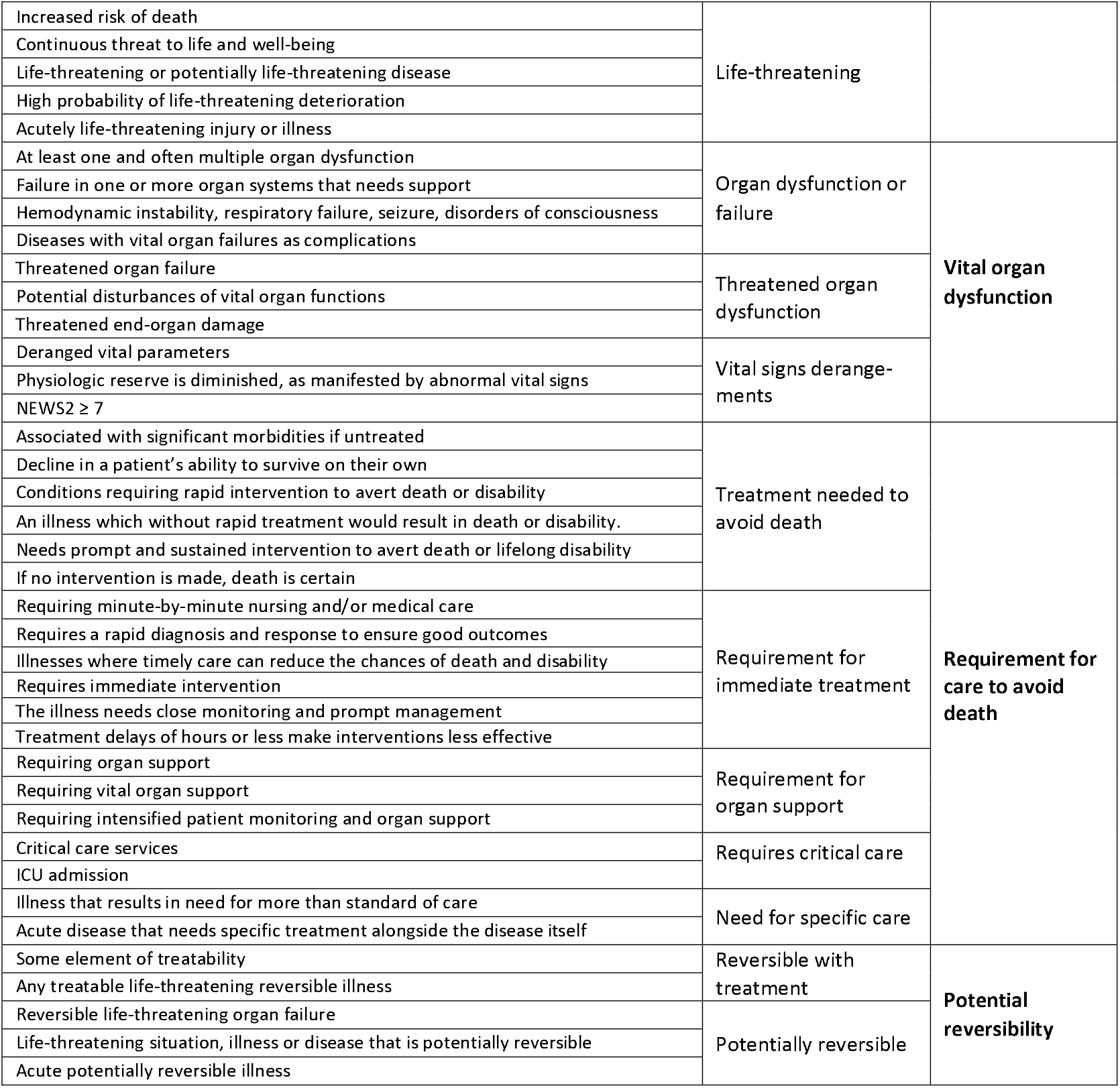
Content analysis for the concept *critical illness*.

**Figure 1:**
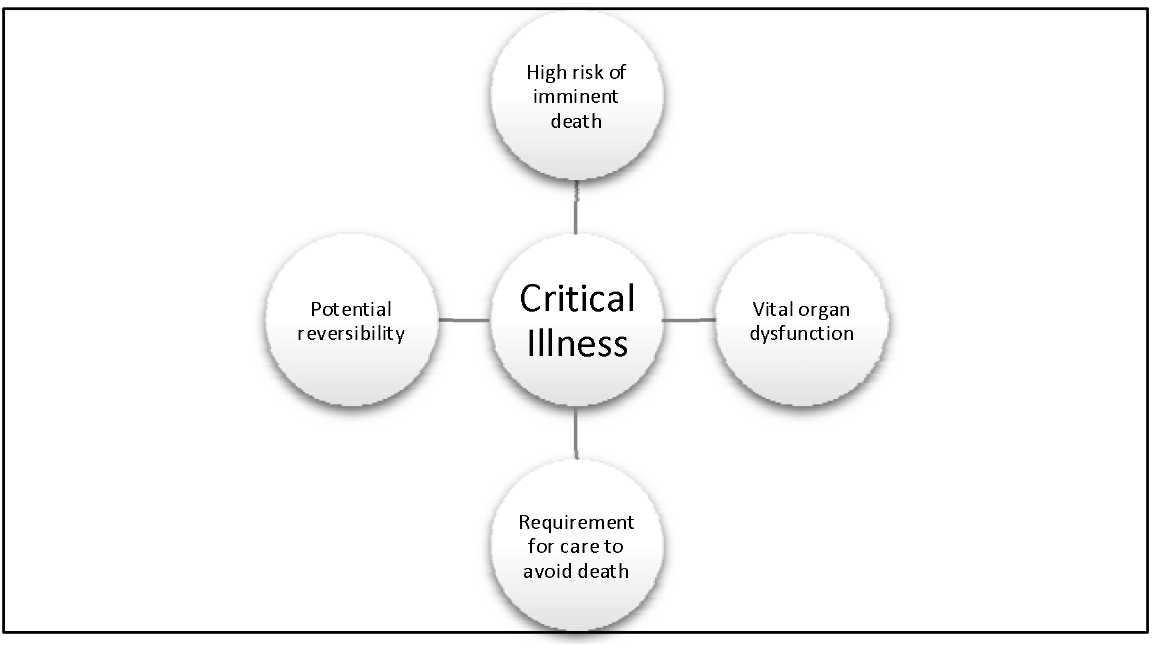
The defining attributes of critical illness.

#### Proposed operational definition

The proposed definition for critical illness is “*Critical illness is a state of ill health with vital organ dysfunction, a high risk of imminent death if care is not provided and the potential for reversibility*.*”*

### Cases

#### A model case of critical illness (a case including all the defining attributes)

A woman has a viral pneumonia. She is breathless and hypoxic with a low oxygen level in her blood (oxygen saturation) of 74%. Her lungs are dysfunctional, and she has a life-threatening condition that is likely to lead to her death in the next few hours. She requires care to support her lungs (oxygen therapy) and if she receives that care, she has a chance of recovery.

#### A related case for critical illness (a case including some of the defining attributes but not the attribute of “imminently life-threatening”)

A man has a chest infection. He has a fever, is coughing up green sputum and feels short-of-breath when walking. He has an oxygen saturation of 91%. He has a serious condition, but it is not imminently life-threatening. He requires treatment, likely with antibiotics, but it is uncertain whether he requires any organ support such as oxygen. His condition is potentially reversible, and he can recover.

#### A contrary case for critical illness (a clear example of “not the concept”)

A woman has lung cancer. She is coughing up small amounts of blood but is able to walk to the hospital. She has an oxygen saturation of 94%. She is sick and she requires treatment. However, her illness is not imminently life-threatening, she has no dysfunctional vital organ and she does not require immediate care. Her condition may or may not be reversible.

#### Antecedents and consequences of Critical Illness

The antecedents of critical illness are the onset of illness, in mild or moderate form, with progressing severity. The consequences of critical illness are either recovery or death.

#### Empirical Referents

There are an estimated 30-45 million cases of critical illness globally each year(1). Many patients are cared for in hospitals with illnesses that are causing vital organ dysfunction and that are imminently life-threatening. There is much work done to identify patients with critical illness such as the use of single severely deranged vital signs(13), or compound scoring systems such as the National Early Warning Score (NEWS) and The Sequential Organ Failure Assessment score (SOFA) (14,15). In hospitals, the severity of patients’ conditions can be assessed using tools such as the Acute Physiology and Chronic Health Evaluation (APACHE) (16) and the Simplified Acute Physiology Score (SAPS)(17).

### Critical Care

#### Defining attributes

A total of 60 codes were identified from the definitions of critical care from the scoping review and expert survey. The codes were analysed into 13 categories and 5 themes. (Table 3) The themes represent the concept’s defining attributes: *identification, monitoring and treatment of critical illness*; vital *organ support*; *initial and sustained care; any care of critical illness*; and *specialized human and physical resources*. (Figure 2)

**Table 3:**
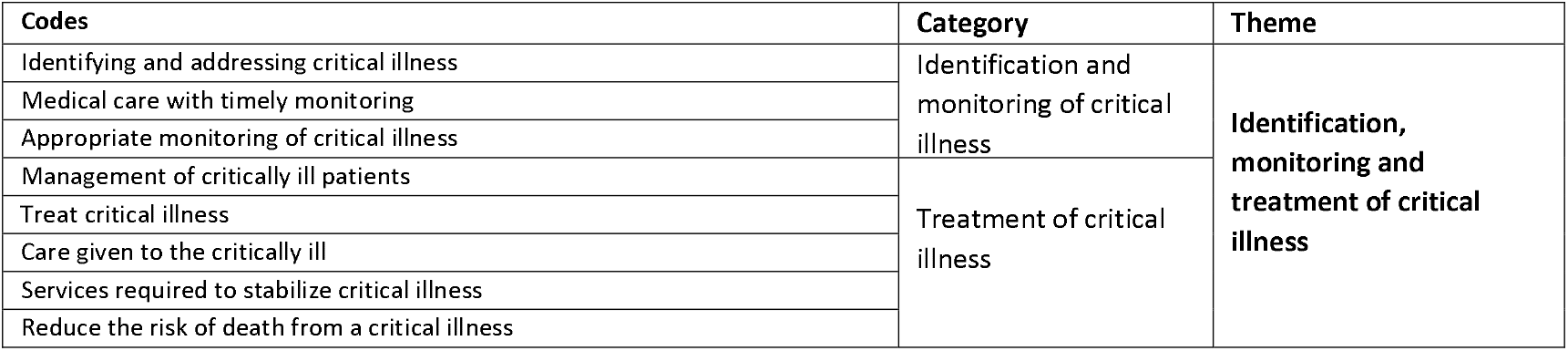
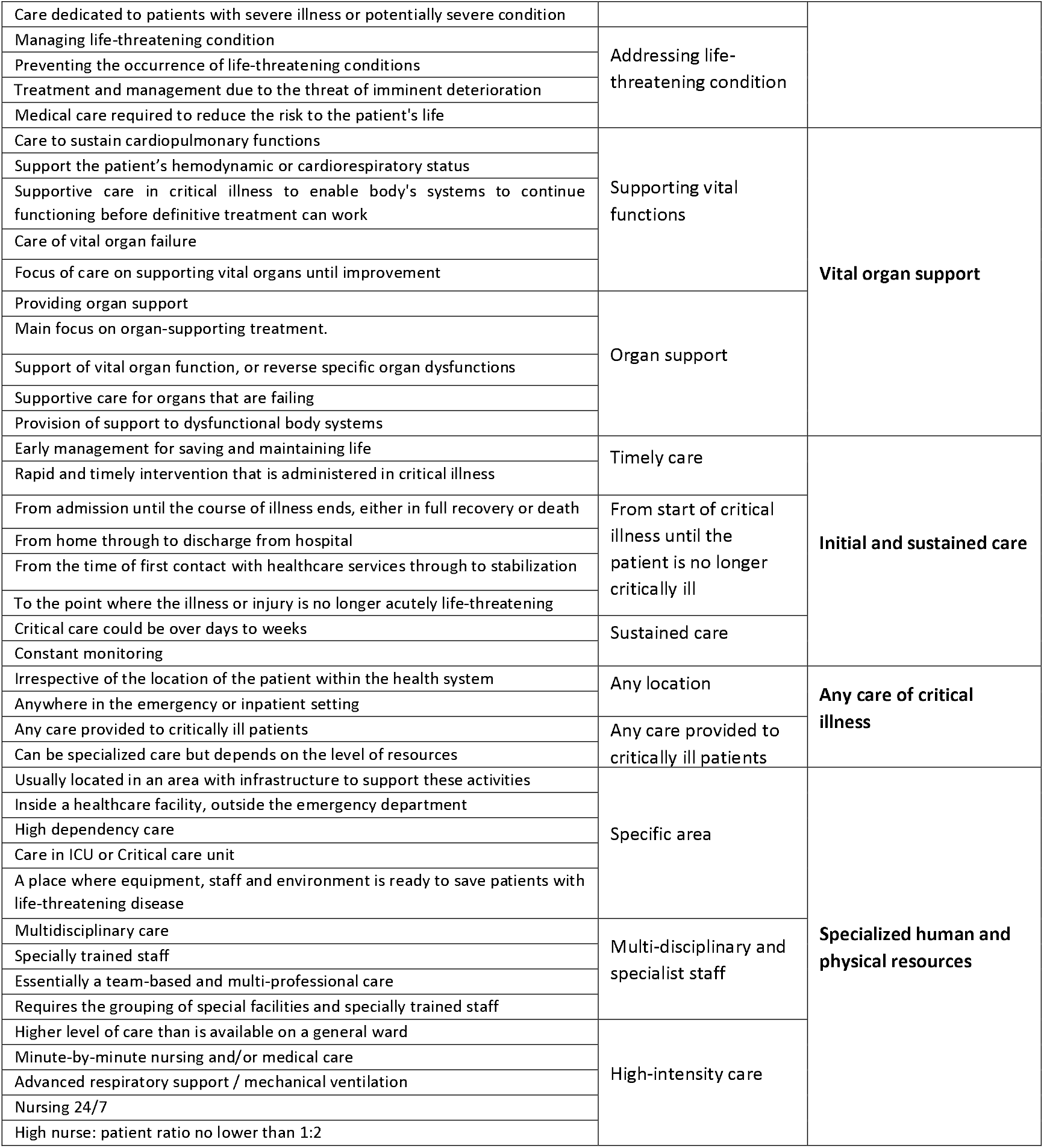
Content analysis for the concept *critical care*.

**Figure 2:**
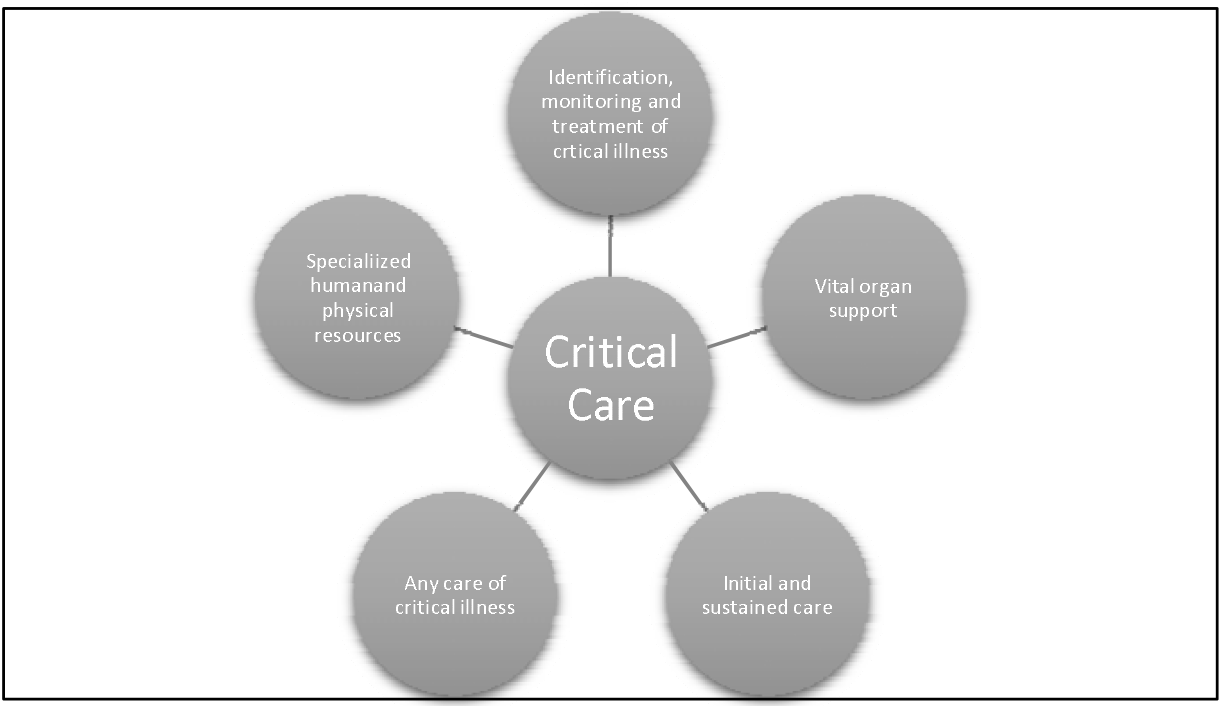
The defining attributes of critical care.

#### Proposed operational definition of *Critical care*

The proposed definition for critical care is “*Critical care is the identification, monitoring and treatment of patients with critical illness through the initial and sustained support of vital organ functions*.*”*

### Cases

#### A model case of critical care (a case including all the defining attributes)

A woman with a viral pneumonia is rapidly identified as critically ill when she arrives at the hospital. She is immediately admitted to a unit with supplies for managing critically ill patients and treatment is started. Nurses and doctors who have been trained in the care of critical illness monitor her regularly, and provide continuous care, titrating the treatments as needed. Continuous oxygen therapy is provided for her life-threatening hypoxia, supporting her respiratory dysfunction, until she has recovered and is no longer critically ill.

#### A related case of critical care (a case including some of the defining attributes but not the attribute of “vital organ support”)

Care in a hospital is provided to a man with a chest infection. A nurse assesses him at arrival to hospital. A doctor admits him to the ward, prescribes antibiotics and decides he is not critically ill and does not require support for any of his vital organs. After four days the doctor discharge him from hospital.

#### A contrary case of critical care (a clear example of “not the concept”)

In the outpatient department, care is provided to a woman with lung cancer. A doctor and a nurse do some investigations and prescribe some medications. She is sent home with a follow-up appointment two weeks later.

#### Antecedents and consequences of critical care

The antecedents of critical care are the contact of the patient with the healthcare system and may include other care of a patient who has not deteriorated to the point of becoming critically ill. The consequences of critical care are either the patient’s recovery or death.

#### Empirical Referents

Many hospitals have wards or units for the provision of critical care, such as Emergency Units, High Dependency Units or Intensive Care Units (ICUs) (18). Critical care can also be provided in general wards, and a recent global consensus specified the care that should be included for all patients with critical illness in any hospital location (19). Rapid Response Teams or Medical Emergency Teams have been introduced into some hospitals, often consisting of staff from the ICU responding to calls from the wards when a critically ill patient has been identified, and providing either critical care on the ward, or transferring the patient to the ICU (20).

## Discussion

We have described how the concepts *critical illness* and *critical care* are used and defined in the literature and by global experts using a concept analysis approach.

Our proposed definition for critical illness of, “*a state of ill health with vital organ dysfunction, a high risk of imminent death if care is not provided and the potential for reversibility*”, is similar to those in some key publications. Chandrashekar et al state that, *“Critical illness is any condition requiring support of failing vital organ systems without which survival would not be possible”* (21) Painter et al write that, *“A critically ill or injured patient is defined as one who has an illness or injury impairing one or more vital organ systems such that there is a high probability of imminent or life-threatening deterioration in the patient’s condition”*(22). Indeed, we found widespread agreement in the literature and expert sources that critical illness concerns life-threatening illness with organ dysfunction.

However, we found diverse and varied usage of the concept concerning the attribute of reversibility and the interface between critical illness and the natural process of dying. Some uses included only illness that was potentially reversible – these sources regarded that for critical illness there should be a possible chance of recovery. Without this, critical illness would be a concept that encompasses the dying process – everyone would be critically ill immediately before death – which would conflict with many clinical uses and understandings of the term., Others had a wider interpretation including all life-threatening illness and did not include reversibility in the definition as it is difficult to identify in the clinical setting, and the concept risks becoming context dependent, (high-resource interventions may reverse some critical illness which would not be possible in low-resource healthcare). Our iterative content analysis method led to our interpretation that reversibility should be included as one of the defining attributes, and this conclusion should be seen as one possible interpretation that can stimulate further discussion.

It is hoped that the proposed definition of critical illness assists communication in the field. Previously, studies about critical illness have focused on patients in certain hospital units, or with diseases or syndromes as proxies for critical illness that exclude some critically ill patients.(1) Our definition of critical illness is not diagnosis or syndrome specific and can be due to any underlying condition. The definition could facilitate the specification of clinical criteria for the identification of critical illness, estimates of the overall burden of critical illness, assessments of outcomes for patients with critical illness across centres and settings, and interventions to improve outcomes.

For critical care, there was greater diversity around its use and definition. There was widespread agreement that critical care is the care of critically ill patients including the support of vital organs. However, there were differing uses around the location of the care and the need for specialized resources. Some sources considered critical care to be only the care provided in certain locations, (such as ICUs or critical care units), or to be care that is always highly specialized or resource-intensive. The World Federation of Societies of Intensive and Critical Care Medicine have suggested that critical care is synonymous with intensive care and is, “*a multidisciplinary and interprofessional specialty dedicated to the comprehensive management of patients having, or at risk of developing, acute, life-threatening organ dysfunction. [Critical care] uses an array of technologies that provide support of failing organ systems, particularly the lungs, cardiovascular system, and kidneys*.”(18) In contrast, other sources used critical care to be inclusive of any care for patients with critical illness, irrespective of location or resources. The Joint Faculty of Intensive Care Medicine of Ireland state that critical care units are those that, “*provide life sustaining treatment for critically ill patients with acute organ dysfunction due to potentially reversible disease*”,(23) and in Belgium, critical care beds have been defined as any beds “*for patients with one or more organ functions compromised*”(24) Hirshon et al strike a balance between these two contrasting views, “*[Critical care is] the specialized care of patients whose conditions are life-threatening and who require comprehensive care and constant monitoring, usually in intensive care units*.” (25)

Our proposed definition of, “*the identification, monitoring and treatment of patients with critical illness through the initial and sustained support of vital organ functions*”, aims to be inclusive. Critical care may include the use of specialized resources but it is not a requirement. We see this as a strength in the definition, as it maintains a patient-centred rather than setting-dependent focus. Critical care when defined in this way can be provided anywhere and does not have to be resource-intensive – it includes both high-resource care in ICUs and lower resource care in other settings. Indeed, critical care can even be provided in general wards, in small health facilities, in the community or in ambulances. High-resource intensive care may not be possible in low-resource settings, but such settings care for many critically ill patients who require critical care(4,26,27). The definition focuses on supporting vital organ functions, emphasising that critical care’s primary focus is treating the critical condition of the patient rather than definitive care for the underlying condition(28,29). Critical care, as we have defined it, can be seen as a system of care of patients with critical illness throughout the course of their illness, from the time of their first contact with healthcare through to resolution of the critical illness or death. Critical care is part of the wider concept of acute care which also includes prehospital care, emergency care, trauma and surgery care, as well as in-patient care in medical, surgical, pediatric, obstetric and other wards(29).

### Strengths and Limitations

To our knowledge, this is the first study attempting to describe the uses and definitions of the concepts *critical illness* and *critical care*, and to identify the defining attributes leading to proposed definitions of the concepts. A strength is the use of both a scoping review of the literature and the inclusion of over one hundred clinical experts as sources. The findings of the analysis should be seen as a first step and we recognise that the use of concepts is fluid and changes over time (6). We were limited to including literature in English and to published studies and guidelines and we may have missed relevant publications in other languages or in other grey literature. Our sample of experts was purposively selected and had global representation but was not perfectly symmetrical to continents, specialty, cadre or gender and we are likely to have missed experts who could have provided valuable contributions. We acknowledge that the proposed definitions may not be universally accepted, and we hope our analysis and findings move the conversation forwards, providing input about how to communicate and collaborate around these vitally important concepts, and ultimately how to improve the care and outcomes for critically ill patients.

## Conclusion

The concepts critical illness and critical care lack consensus definitions and have varied uses. Through concept analysis of the uses in the literature and among experts we propose the definitions: “*Critical illness is a state of ill health with vital organ dysfunction, a high risk of imminent death if care is not provided and the potential for reversibility*” and “*Critical care is the identification, monitoring and treatment of patients with critical illness through the initial and sustained support of vital organ functions*.”

## Data Availability

The study data are available from the corresponding author on reasonable request

## Acknowledgements

We thank all the experts who participated in the study.

## Author Contributions

TB & OS designed the study. RKK, TT, HM and TB collected the data. All the authors contributed to analysing the data. RKK and TB wrote the first draft of the manuscript. All authors critically reviewed the manuscript and approved the final version. The corresponding author attests that all listed authors meet authorship criteria and that no others meeting the criteria have been omitted.

## Disclaimer

We confirm the independence of researchers and that all authors in study can take responsibility for the integrity of the data and the accuracy of the data analysis.

## Patient Consent for Publication

Not required

## Ethics Approval

The Research Ethics Committee of the London School of Hygiene and Tropical Medicine approved the study (Reference number 22661).

## Provenance and Peer Review

Not commissioned, externally reviewed

## Data Availability Statement

The study data are available from the corresponding author on reasonable request

## Supplementary Files

Supplementary Tables 1 and 2

